# Federal Vaccine Policy and Interstate Variation in COVID-19 Vaccine Coverage in India

**DOI:** 10.1101/2021.08.16.21262113

**Authors:** Kanchan Mukherjee

## Abstract

**Introduction:** On August 13, 2021, India completed 30 weeks of vaccination against COVID-19 for its eligible citizens. While the vaccination has made progress, there has been no study analyzing the federal/union vaccine policy and its effect on vaccination coverage across Indian states. In this context, this study analyses the federal vaccination policy and its effect on interstate variation in vaccine coverage and the correlation of state economy with vaccination coverage.

**Methods:** The study analyses vaccine policy documents, secondary data on vaccination coverage and state gross domestic product (GDP) available in public domain. ANOVA test has been used to assess the effect of vaccine policy on interstate vaccine coverage and correlation-regression analysis has been conducted to assess the type and strength of association between gross state domestic product and vaccination coverage.

**Results:** Interstate variation in vaccination coverage in the first 15 weeks was the least (F=3.5), when vaccine procurement and supply was entirely provided by the union/federal government and vaccination was limited to priority groups. However, with the extension of vaccine policy to other groups and reduction in federal government involvement in vaccine procurement, the interstate variation in vaccination coverage increased significantly (F=10.74) by the end of 30 weeks. The highest interstate variation was observed in the period between 23-30 weeks (F=25.31). State GDP was positively and strongly correlated with state vaccination coverage with a high coefficient of correlation (R=0.94) and high coefficient of determination (R^2^= 0.88).

**Conclusions:** The study finds that federal procurement and supply of vaccination among prioritized groups has been the best strategy till date to address the inequity in vaccination coverage across the states of India.

## Introduction

The largest national COVID-19 vaccination programme in the world began in India on January 16, 2021 (Kumar et al., 2021). Apart from utilizing the existing human resources and cold-chain infrastructure, this vaccine drive has also given impetus to the country’s vaccine manufacturing capacity. COVID-19 vaccination in the country commenced with vaccination to all healthcare workers. The program was expanded with time to include vaccination of front line workers, citizens more than 60 years of age, citizens more than 45 years of age and eventually citizens more than 18 years of age. While the huge eligible population and increase in cases in India has affected the availability of vaccine, as on July 31, 2021 India had completed administering 461.5 million doses (PIB, July 31, 2021), which was much higher than the target proposed of 300 million doses (Padma, 2021). However, there are reports that vaccine doses administered have been uneven over the country across states and districts. Also, federal vaccine policy guidelines for states have changed with time. Initially (first 15 weeks), all vaccines were procured and supplied by federal government after which the state governments and private hospitals were given the authority to procure vaccines directly (MOHFW, 2021). Hence, this study proposes to assess the effect of the change in federal vaccine policy on the interstate and intrastate vaccination coverage with time. Since, procurement of vaccines, administration, logistics etc. could be affected by the level of state’s economic development, the study also analysed the correlation between State Gross Domestic Product (GDP) and vaccine doses administered among these states during this time period. Hence, this article answers the following research questions:

1. Is there a significant interstate and intrastate variation in vaccine coverage with time?
2. What has been the effect of the change in federal vaccination policy to these variations?
3. What is the correlation of state GDP with the state vaccine coverage?

## Methods

This study is based entirely on secondary data available in the public domain. For the data on vaccination doses given, the Co-WIN dashboard (https://dashboard.cowin.gov.in/) has been used as a source of data. For the State GDP data, the information from ministry of statistics and programme implementation available at statisticstimes.com has been used. Based on the Ministry of Health and Family Welfare (MOHFW) website data, the ten states worst affected by COVID-19 during the vaccination period till date, have been selected. Documents on vaccine policy guidelines issued by the union/federal government to the state governments have been reviewed to analyse the vaccination policy and federal-state vaccine procurement strategy over time. ANOVA was applied to test the interstate variation in vaccine coverage with time and correlation-regression analysis was used to test the association of state GDP with vaccine coverage. The data entry and analysis was performed using Excel.

## Results

India completed 30 weeks of vaccination on August 13, 2021, by which time a total of 530 million doses were administered (PIB, August 13, 2021). Under the National COVID Vaccination Program (MOHFW, 2021), from 16th January to 30th April 2021, 100% of vaccine doses were procured by Government of India and provided free of cost to state governments. During this period, the federal government received suggestions by many state governments to be permitted the flexibility to procure vaccine directly and administer them as per their own prioritization based on local requirements. Based on this request and effective from 1st May, 2021, the first union/federal COVID-19 vaccine policy change was made. As per this revised vaccine policy, the federal government would procure 50% of the vaccine produced and continue to provide them to states free of cost for administering to priority groups while the state government and private hospitals were empowered to directly procure from the remaining 50% vaccine pool. However, many states and smaller and remoter private hospitals faced difficulties in managing the funding, procurement and logistics of vaccines. Hence, the guidelines were revised again with effect from June 21, 2020, wherein it was decided that the federal government would procure 75% of vaccines required for the states, while states and private hospitals could directly procure from among the remaining 25% required (MOHFW, 2021). Based on the above changes in federal vaccine policy, the 30-week period could be divided as follows:

1. First 15 weeks of vaccination: 100% federal government funded vaccines.
2. Weeks 16-22: 50% federal funded vaccines
3. Weeks 23-30: 75% federal funded vaccines

The variation in COVID-19 vaccine coverage in terms of doses given across the 10 worst affected states during the 30 weeks of vaccination is shown in Figure 1.

**Figure 1.**
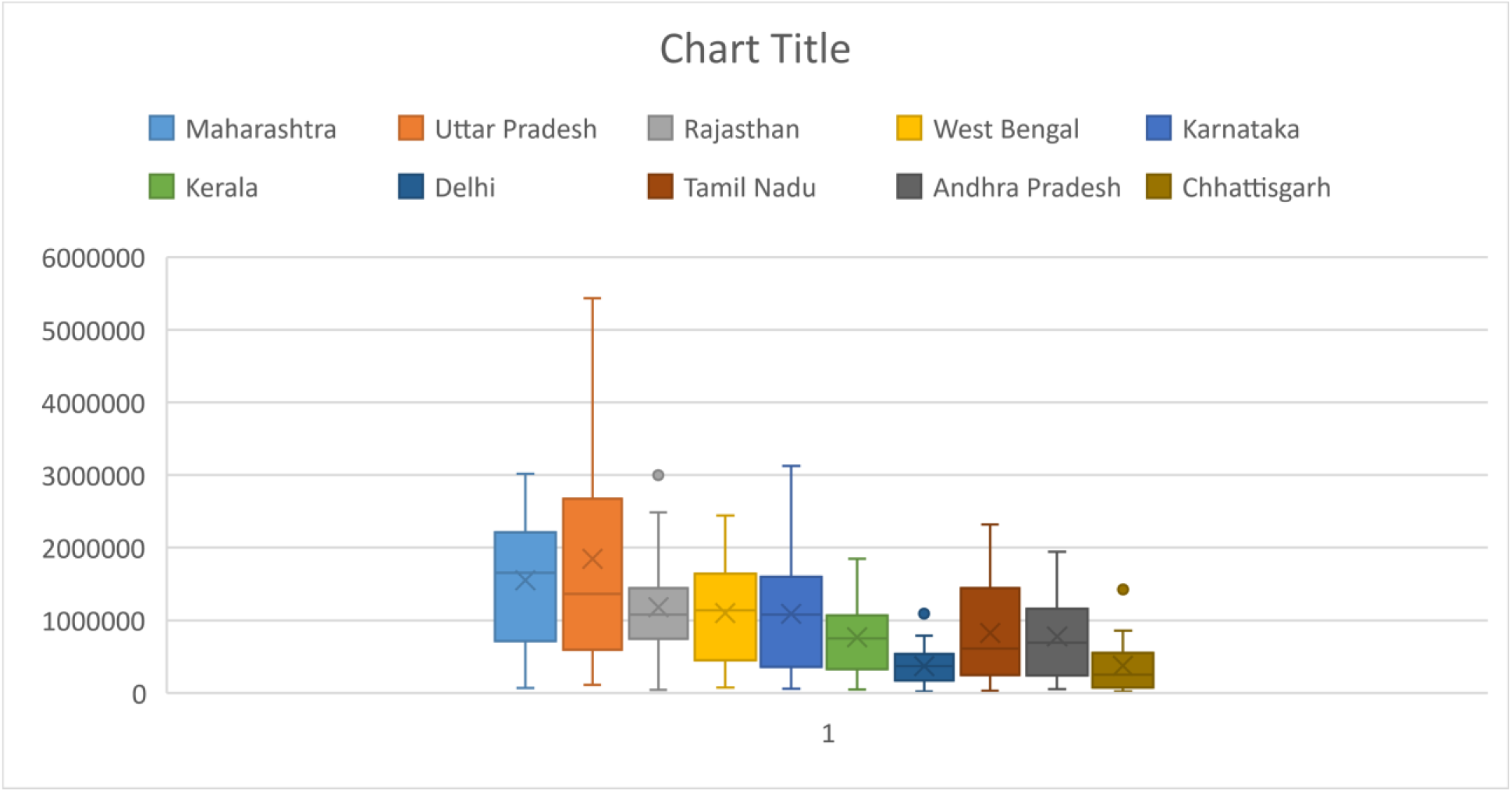
Variation in vaccination coverage among 10 worst affected states in 30 weeks

To test the significance level of the above variations, analysis of variance (ANOVA) test was used to understand the variance in vaccine doses across these 10 states during the 30-week period. The ANOVA results (Table 1) show a significant variation in vaccination coverage with time within states as well as between states. There was statistically highly significant (p = 2.15 × 10^−14^) interstate differences in the vaccination coverage in these 30 weeks of vaccination (F=10.74).

**Table 1.** ANOVA results of interstate and intrastate differences in 30 weeks of vaccination

| *States* | *Weeks* | *Vaccine doses* | *Average* | *Variance* |
| --- | --- | --- | --- | --- |
| Maharashtra | 30 | 46548761 | 1551625 | 8.70E+11 |
| Uttar Pradesh | 30 | 55458462 | 1848615 | 2.31E+12 |
| Rajasthan | 30 | 35493110 | 1183104 | 6.60E+11 |
| West Bengal | 30 | 32953291 | 1098443 | 4.49E+11 |
| Karnataka | 30 | 32617293 | 1087243 | 5.68E+11 |
| Kerala | 30 | 23008463 | 766949 | 2.76E+11 |
| Delhi | 30 | 11144261 | 371475 | 6.30E+10 |
| Tamil Nadu | 30 | 24833682 | 827789 | 4.69E+11 |
| Andhra Pradesh | 30 | 23319070 | 777302 | 3.62E+11 |
| Chhattisgarh | 30 | 11337355 | 377912 | 1.47E+11 |

| ANOVA |  |  |  |  |  |  |
| --- | --- | --- | --- | --- | --- | --- |
| *Source of Variation* | *SS* | *df* | *MS* | *F* | *P-value* | *F crit* |
| Between Groups | 5.97E+13 | 9 | 6.6322E+12 | 10.74 | 2.15E-14 | 1.91 |
| Within Groups | 1.79E+14 | 290 | 6.1759E+11 |  |  |  |
| Total | 2.39E+14 | 299 |  |  |  |  |

To assess whether the vaccine policy had any effect on these variations, the study using ANOVA test first compared the F value for two time periods: weeks 1-15 (full federal procurement) versus weeks 16-30 (partial federal procurement). ANOVA results of weeks 1-15 and weeks 16-30 are shown in table 2 and 3 respectively. The F value for interstate variation in vaccination coverage during full federal vaccine procurement policy was 3.5 (Table 2) while it was 15.79 (Table 3) during the partial federal procurement policy indicating a much higher interstate variation when the vaccine policy was changed to partial federal procurement. While both the interstate variations were significant at p<0.05, the p value of the interstate variation with partial federal procurement was significantly much lower (p=1.32 × 10^−14^) (Table 3).

**Table 2.** ANOVA results first 15 weeks (100 % Federal funded and procured vaccines)

| *States* | *Weeks* | *Vaccine doses* | *Average* | *Variance* |
| --- | --- | --- | --- | --- |
| Maharashtra | 15 | 15618307 | 1041220 | 7.92E+11 |
| Uttar Pradesh | 15 | 11995025 | 799668 | 3.97E+11 |
| Rajasthan | 15 | 12543605 | 836240 | 6.08E+11 |
| West Bengal | 15 | 10775800 | 718387 | 3.06E+11 |
| Karnataka | 15 | 9231248 | 615417 | 2.79E+11 |
| Kerala | 15 | 7207832 | 480522 | 1.35E+11 |
| Delhi | 15 | 3180770 | 212051 | 2.94E+10 |
| Tamil Nadu | 15 | 5550730 | 370049 | 9.52E+10 |
| Andhra Pradesh | 15 | 6220545 | 414703 | 1.18E+11 |
| Chhattisgarh | 15 | 5418687 | 361246 | 1.54E+11 |

ANOVA
| *Source of Variation* | *SS* | *df* | *MS* | *F* | *P-value* | *F crit* |
| --- | --- | --- | --- | --- | --- | --- |
| Between Groups | 9.17E+12 | 9 | 1.02E+12 | 3.50 | 0.0006 | 1.95 |
| Within Groups | 4.08E+13 | 140 | 2.91E+11 |  |  |  |
| Total | 5E+13 | 149 |  |  |  |  |

**Table 3.** ANOVA results for weeks 16-30 (Partial federal procurement)

| *States* | *Weeks* | *Vaccine doses* | *Average* | *Variance* |
| --- | --- | --- | --- | --- |
| Maharashtra | 15 | 30930454 | 2062030 | 4.51E+11 |
| Uttar Pradesh | 15 | 43463437 | 2897562 | 2.04E+12 |
| Rajasthan | 15 | 22949505 | 1529967 | 5.01E+11 |
| West Bengal | 15 | 22177491 | 1478499 | 3.14E+11 |
| Karnataka | 15 | 23386045 | 1559070 | 4.21E+11 |
| Kerala | 15 | 15800631 | 1053375 | 2.61E+11 |
| Delhi | 15 | 7963491 | 530899 | 4.66E+10 |
| Tamil Nadu | 15 | 19282952 | 1285530 | 4.27E+11 |
| Andhra Pradesh | 15 | 17098525 | 1139902 | 3.5E+11 |
| Chhattisgarh | 15 | 5918668 | 394578 | 1.5E+11 |

ANOVA
| *Source of Variation* | *SS* | *df* | *MS* | *F* | *P-value* | *F crit* |
| --- | --- | --- | --- | --- | --- | --- |
| Between Groups | 7.04E+13 | 9 | 7.82663E+12 | 15.79 | 1.32E-17 | 1.95 |
| Within Groups | 6.94E+13 | 140 | 4.95713E+11 |  |  |  |
| Total | 1.4E+14 | 149 |  |  |  |  |

The above results also reflect the variation in capacity among state governments to manage the vaccination coverage challenge. This variation in capacity could be affected by the economic condition of the state among other factors and hence, a scatter plot (Figure 2) and correlation-regression analysis (Table 4) was used to understand the correlation of state GDP with vaccination coverage across these states for weeks 16-30.

**Table 4.**
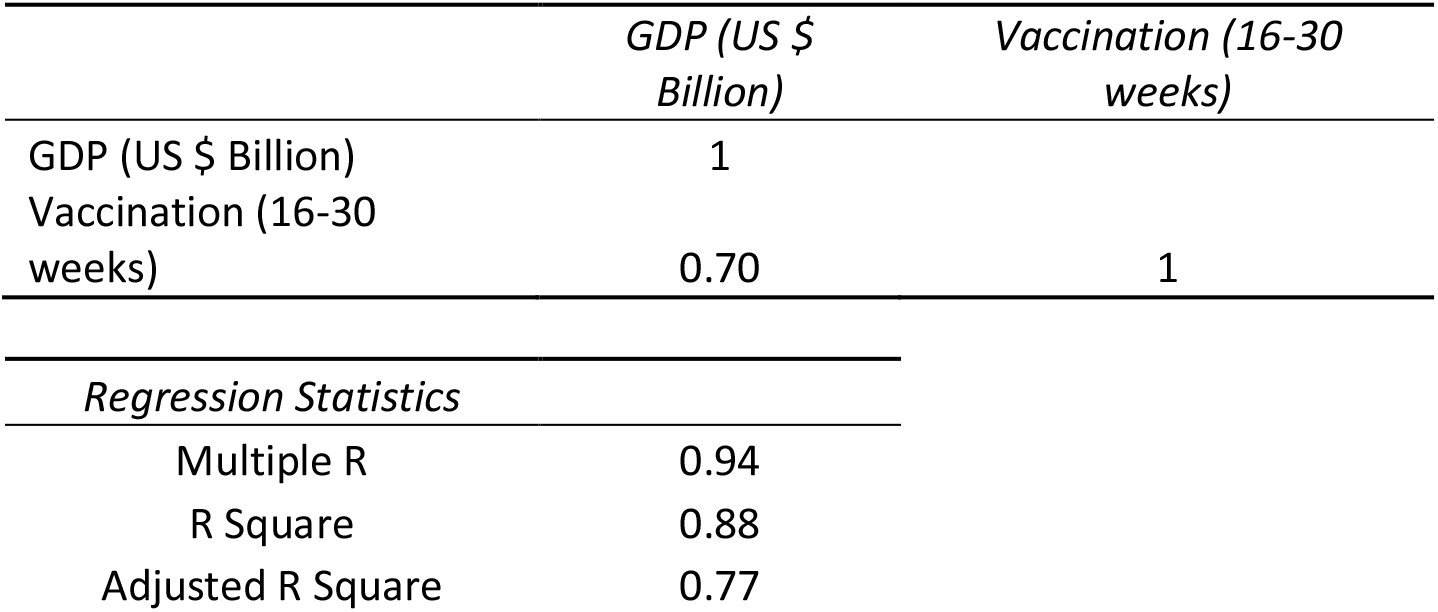
Correlation regression analysis of state GDP and vaccination coverage

**Figure 2.**
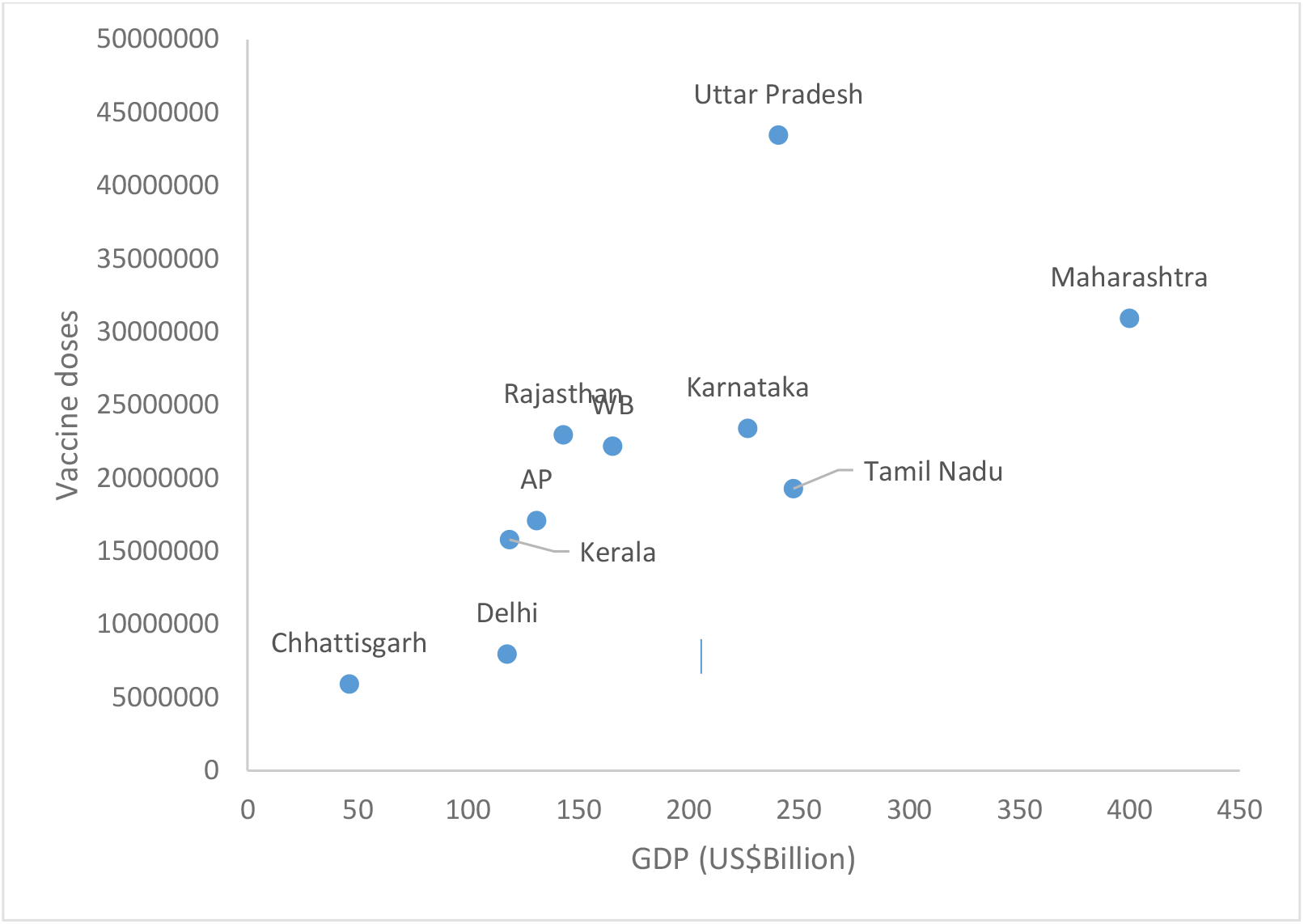
Scatter plot of state GDP and vaccination doses

Correlation analysis shows a positive association between state GDP and vaccination coverage. The coefficient of determination R^2^ value of 0.88 and adjusted R^2^ of 0.77 indicates a strong correlation between state GDP and vaccine doses administered. 88% of the variation in vaccine coverage can be explained by variation in state GDP. Also the coefficient of correlation R is 0.94, indicating a strong and positive relationship between state GDP and vaccine coverage. Hence, state GDP is strongly correlated with vaccine coverage.

Since, the vaccination policy changed the second time in week 23, disaggregated ANOVA analysis was used to compare interstate variation in vaccination coverage between weeks 16-22 and weeks 23-30. Table 5 and 6 shows the ANOVA results for these time periods.

**Table 5.**
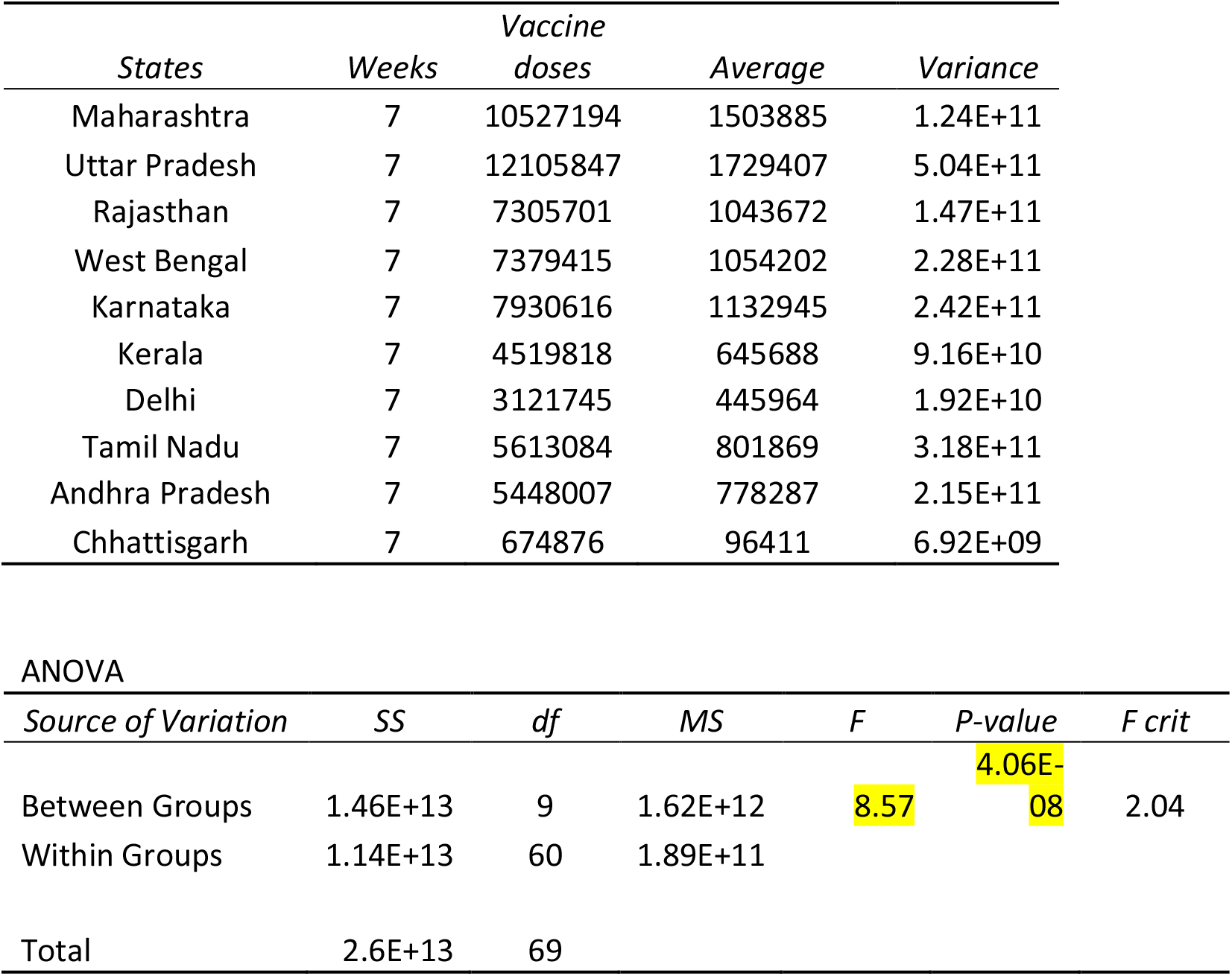
Weeks 16-22 (50% federal procured vaccines and 50% state/private procurement)

| *States* | *Weeks* | *Vaccine doses* | *Average* | *Variance* |
| --- | --- | --- | --- | --- |
| Maharashtra | 7 | 10527194 | 1503885 | 1.24E+11 |
| Uttar Pradesh | 7 | 12105847 | 1729407 | 5.04E+11 |
| Rajasthan | 7 | 7305701 | 1043672 | 1.47E+11 |
| West Bengal | 7 | 7379415 | 1054202 | 2.28E+11 |
| Karnataka | 7 | 7930616 | 1132945 | 2.42E+11 |
| Kerala | 7 | 4519818 | 645688 | 9.16E+10 |
| Delhi | 7 | 3121745 | 445964 | 1.92E+10 |
| Tamil Nadu | 7 | 5613084 | 801869 | 3.18E+11 |
| Andhra Pradesh | 7 | 5448007 | 778287 | 2.15E+11 |
| Chhattisgarh | 7 | 674876 | 96411 | 6.92E+09 |

| ANOVA |  |  |  |  |  |  |
| --- | --- | --- | --- | --- | --- | --- |
| *Source of Variation* | *SS* | *df* | *MS* | *F* | *P-value* | *F crit* |
| Between Groups | 1.46E+13 | 9 | 1.62E+12 | 8.57 | 4.06E-08 | 2.04 |
| Within Groups | 1.14E+13 | 60 | 1.89E+11 |  |  |  |
| Total | 2.6E+13 | 69 |  |  |  |  |

**Table 6.** ANOVA results for weeks 23-30 (75% federal procured vaccines and 25% state/private procurement)

| *States* | *Weeks* | *Vaccine doses* | *Average* | *Variance* |
| --- | --- | --- | --- | --- |
| Maharashtra | 8 | 20403260 | 2550408 | 2.12E+11 |
| Uttar Pradesh | 8 | 31357590 | 3919699 | 1.08E+12 |
| Rajasthan | 8 | 15643804 | 1955476 | 4.33E+11 |
| West Bengal | 8 | 14798076 | 1849760 | 9.52E+10 |
| Karnataka | 8 | 15455429 | 1931929 | 2.95E+11 |
| Kerala | 8 | 11280813 | 1410102 | 1.32E+11 |
| Delhi | 8 | 4841746 | 605218 | 6.32E+10 |
| Tamil Nadu | 8 | 13669868 | 1708734 | 1.43E+11 |
| Andhra Pradesh | 8 | 11650518 | 1456315 | 2.71E+11 |
| Chhattisgarh | 8 | 5243792 | 655474 | 1.27E+11 |

| ANOVA |  |  |  |  |  |  |
| --- | --- | --- | --- | --- | --- | --- |
| *Source of Variation* | *SS* | *df* | *MS* | *F* | *P-value* | *F crit* |
| Between Groups | 6.49E+13 | 9 | 7.2E+12 | 25.31 | 1.08E-18 | 2.02 |
| Within Groups | 2E+13 | 70 | 2.9E+11 |  |  |  |
| Total | 8.49E+13 | 79 |  |  |  |  |

Comparing the F score across the three time periods emerging from the change in federal vaccine policy, it is evident that the interstate variation in vaccine coverage has increased significantly with time (Table 7).

**Table 7.** Effect of federal vaccine policy change on interstate state vaccine coverage variation

| Vaccine Policy | F value | P value |
| --- | --- | --- |
| 100% Federal procurement policy (15 weeks) [only priority groups eligible for vaccination] | 3.50 (lowest variation) | $6 \times 10^{-4}$ |
| 50% federal procurement policy (7 weeks) | 8.57 | $4.06 \times 10^{-8}$ |
| 75% federal procurement policy (8 weeks) | 25.31 (maximum variation) | $1.08 \times 10^{-18}$ |

## Conclusions

There are statistically significant interstate differences and intrastate differences with time in the vaccine coverage in the first 30 weeks of vaccination. The first 15 weeks showed the least variation when the vaccine procurement and supply was fully with the federal government. This was also the period when vaccination was restricted to priority groups and did not include the 18-44 years’ age group for vaccination. From May 1, 2021 onwards (week 16), with the 18-44 years’ age group being eligible for vaccines and the federal government decreasing its role as vaccine supplier from 100% to 50%, the demand overtook the supply in a big way. This resulted in a significant increase in interstate variations in vaccination coverage which could be explained by the state’s economic status as the correlation-regression analysis found a significantly strong association with state GDP and vaccination coverage. This interstate variation has only increased with time, inspite of the federal government increasing its role in vaccine supply after June 21, 2021 from 50%-75%. This is probably a reflection of the extent of vaccine shortage in this period and the federal government is itself finding it hard to procure vaccines.

The study findings provide a statistical objective assessment of the effect of changes in federal vaccine policy on intrastate and interstate variation in vaccination coverage in India in the last 30 weeks. The vaccine policies have had significant impact on both procurement as well as demand of vaccines in India in the last 30 weeks. Federal procurement and supply of vaccination among prioritized groups has been the best strategy till date to address the vaccine inequity across the states of India. Since, vaccination coverage is affected significantly by economic status of states, economically weak states are disadvantaged. Given the fact that interstate variations were the least with a 100% federal procurement and supply policy, it may be prudent to retain the 100% federal procurement strategy for a more equitable distribution of vaccine coverage. If the federal government is the sole procurer of vaccine, it allows for a larger and uniform vaccine purchase and supply system, which can be used to distribute vaccines as per requirements in different states and is independent of the state’s economic status.

## Data Availability

All data used for this study is publicly available

